# Magnitude, global variation, and temporal development of the COVID-19 infection fatality burden

**DOI:** 10.1101/2021.12.17.21267986

**Authors:** Christina Bohk-Ewald, Enrique Acosta, Tim Riffe, Christian Dudel, Mikko Myrskylä

## Abstract

How deadly is an infection with SARS-CoV-2 worldwide over time? This information is critical for developing and assessing public health responses on the country and global levels. However, imperfect data have been the most limiting factor for estimating the COVID-19 infection fatality burden during the first year of the pandemic. Here we leverage recently emerged compelling data sources and broadly applicable modeling strategies to estimate the crude infection fatality rate (cIFR) in 77 countries from 28 March 2020 to 31 March 2021, using 2.4 million reported deaths and estimated 435 million infections by age, sex, country, and date. The global average of all cIFR estimates is 1.2% (10th to 90th percentile: 0.2% to 2.4%). The cIFR varies strongly across countries, but little within countries over time, and it is often lower for women than men. Cross-country differences in cIFR are largely driven by the age structures of both the general and the truly infected population. While the broad trends and patterns of the cIFR estimates are more robust, we show that their levels are uncertain and sensitive to input data and modeling choices. In consequence, increased efforts at collecting high-quality data are essential for accurately estimating the cIFR, which is a key indicator for better understanding the health and mortality consequences of this pandemic.

---

How fatal is an infection with SARS-CoV-2? How has the crude infection fatality rate (cIFR) developed over time and what drives its global variation? How large would the cIFR be in response to changes in medical treatment or spread of deadlier SARS-CoV-2 variants? Estimating and analyzing the cIFR in many countries over time is important for managing and learning from this global health crisis.

The most limiting factor for estimating the cIFR—defined as total number of deaths over total number of infections—in countries worldwide over time is data. Next to inadequate testing coverage and test sensitivity, cause-of-death misclassification, and errors and delays in reporting COVID-19-related deaths and cases (1), the numbers of reported deaths have often not been harmonized by age and sex over time (2); and the number of test-positive cases underestimate the total numbers of infected individuals (3). Early but limited data on COVID-19 have allowed to estimate the case fatality rate for some countries (4), but they have been unsuitable to estimate the cIFR for many countries over time—particularly because the size of the truly infected population has been unknown.

Our approach is to leverage recently emerged data sources and modeling strategies to calculate the cIFR, using data on cumulative numbers of reported deaths by age from the COVerAGE database (2) and a demographic scaling model (3) to estimate cumulative numbers of infections by age. This estimation approach uses age-specific IFRs from China (5) as the baseline. It scales them to other countries by assuming that age-specific IFRs vary across countries proportionally to remaining life expectancy. As a general marker of population health, remaining life expectancy captures vulnerability to COVID-19 with respect to population age, pre-existing conditions, and medical services. We also assume that the scaled baseline IFRs by age are constant within countries over time. This assumption is often made (6-8), even though improvement in medical treatment (9), shortages of critical medical equipment, or spread of deadlier SARS-CoV-2 variants (10) may change the age-specific IFRs. We explore the sensitivity of our cIFR estimates to deviations from these key assumptions.

## Results

Figure 1A shows the spatio-temporal distribution of all cIFR estimates, and Fig. 1B shows the set of countries available over time. The cIFR is, on average, 1.2% (10th to 90th percentile: 0.2% to 2.4%), comparatively high in Europe (average 1.7%), Northern America (1.4%), and Australia and New Zealand (2.2%); and low in Africa (0.8%), Asia (0.6%), and Latin America and the Caribbean (0.3%). The cIFR has converged quickly to comparatively stable levels after the outbreak of the pandemic; cross-country variation is large, within-country variation is low. This time stability of the cIFR estimates is driven by the stability of the age distribution of cumulative deaths.

**Figure 1.**
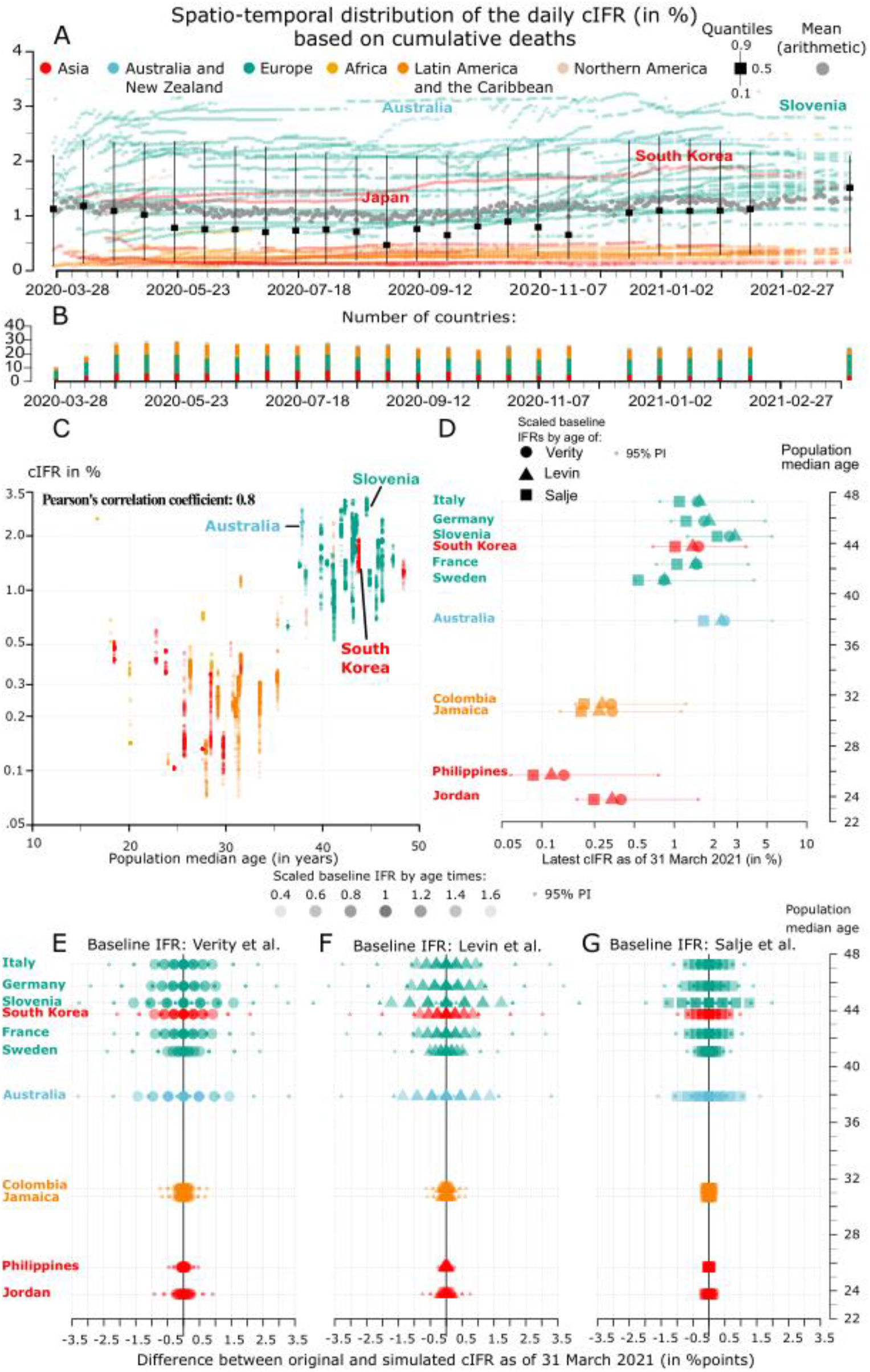
Level, global variation, and temporal development of the cIFR. (A) cIFR by country and date. (B) Number of countries per date. (C) Correlation between log cIFR and population median age. (D) cIFR as of 31 March 2021 based on scaled baseline IFRs of Verity (5), Levin (7), or Salje (12). (E-G) Difference between original and simulated cIFR estimates as of 31 March 2021.

Figure 1C shows the correlation of 0.8 between log of cIFR and population median age, indicating that the older populations of Europe, Australia, New Zealand, and Northern America have higher cIFRs than the younger populations. The log of cIFR also strongly correlates with the median age of the estimated and confirmed infected population: 0.9 for both (11). These findings suggest that the age structures of both the general and the infected population are key drivers of the level and global variation of the cIFR.

Figure 1D shows that the estimated cIFRs as of 31 March 2021 are associated with high levels of uncertainty (average width of the 95% prediction interval is 2.8 percentage points) and sensitive towards baseline IFRs by age (average cIFR varies from 0.9% when based on French data of Salje (12) to 1.3% when based on Chinese data of Verity (5) or data of 28 highly-developed locations of Levin (7)). This analysis suggests that the choice of the baseline IFRs by age (11) does not change the broad patterns and trends of the cIFR across countries, but that it does add to the uncertainty of the level of these estimates within countries.

Figure 1E-G illustrates how the cIFR as of 31 March 2021 would respond to changes in the scaled baseline IFRs by age within selected countries. These scenarios showcase what would happen to the cIFR when, for example, medical treatment was improved (9,13), vaccination was broadly accessible (14), critical medical equipment was running short, or deadlier SARS-CoV-2 variants were spread more widely (10). This simulation also highlights the sensitivity of our original cIFR estimates with respect to potential misestimation of the scaled baseline IFRs by age (3). If the scaled baseline IFRs by age in each country were 60% higher—corresponding to the increased risk of death associated with some new SARS-CoV-2 variants (10)—the cIFR would increase, on average, by 0.8 percentage points across all the countries covered. The effects on cIFR, however, differ strongly by country. For example, the comparatively large impact in Slovenia and Australia stems from their infected populations being estimated to be comparatively old (11). This analysis indicates that the levels, not the patterns and trends, of the cIFR estimates are sensitive towards changes in or misestimation of the level of the scaled IFRs by age.

Figure 2 shows that the cIFR varies by sex, being on average 0.1 percentage points smaller for females than for males. This is not surprising, as the baseline IFRs by age are consistently smaller for women than for men (11,15). European countries, Japan, and Australia have the largest sex differences, with European countries mostly having lower cIFR for women, whereas Australia has higher cIFR for women. This cross-country variation is likely to be related to the age structure of infected female and male population, as well as to sex differences in age-specific IFRs.

**Figure 2.**
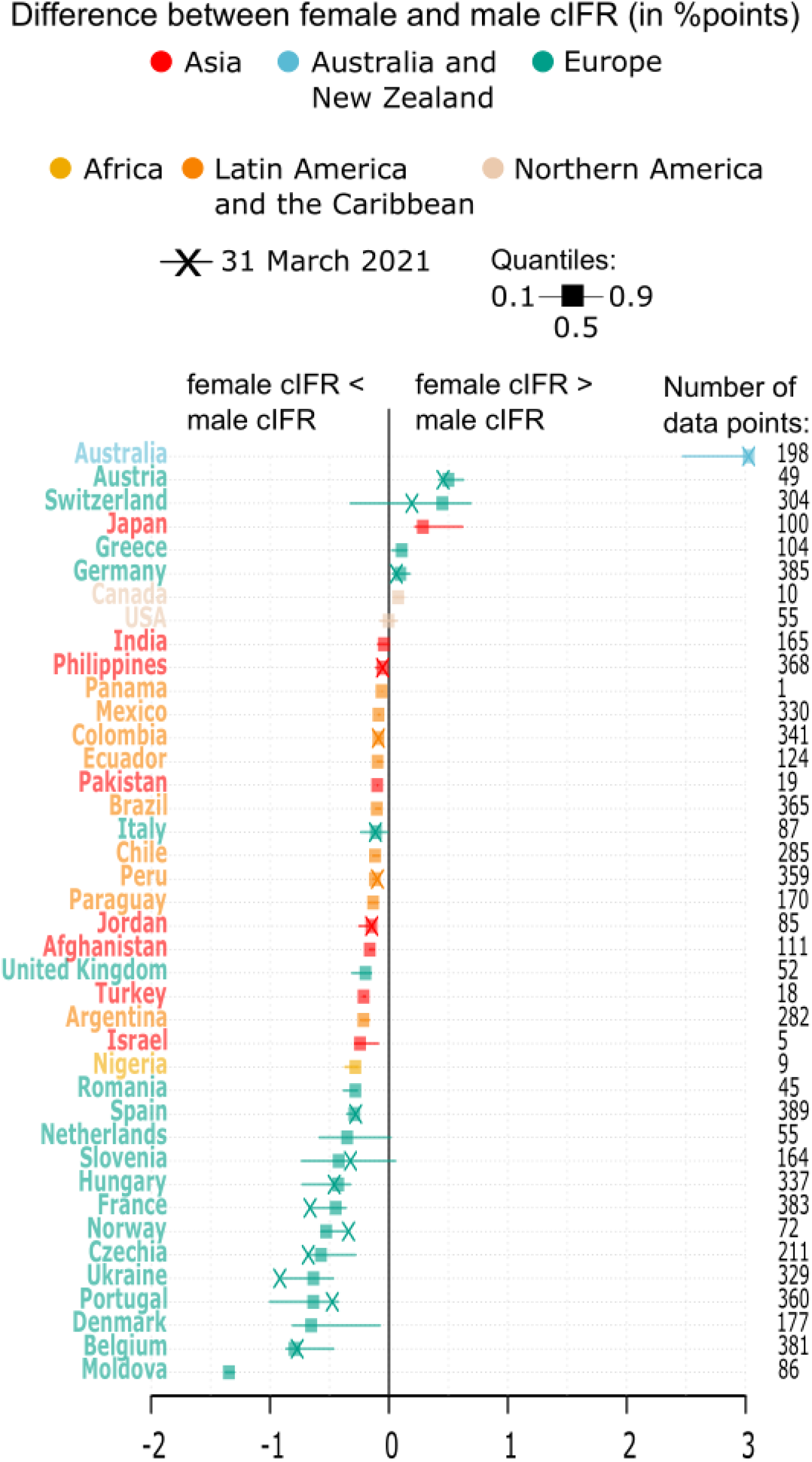
Difference between female and male cIFR.

## Discussion

We use best available data (2) and recently emerged methods that avoid important biases in the infected population (3) to estimate the cIFR. Despite this, the data is imperfect and modeling assumptions are strong. Therefore, we have focused on interpreting the broad patterns and trends of the cIFR that are likely to be robust to systematic bias. We also have attempted to gauge the uncertainty of our cIFR estimates by analyzing their sensitivity to input data from various sources: using (i) three different baseline IFRs by age (5,7,11,12), (ii) reported or estimated excess deaths (2,11), and (iii) estimated or confirmed cases (2,3,11). These sensitivity analyses suggest that the broad patterns reported in this manuscript are robust to changes (i)-(iii), but the overall estimated level of cIFR varies depending on the data and modeling choice.

Our study has enabled us to draw a landscape of the level, global variation, and temporal development of the COVID-19 infection fatality burden during the first year of the coronavirus pandemic. It reveals that cIFR has converged to quite stable levels within and between countries over time and is often larger for men than women. Our analysis suggests that the age structures of both the general and the truly infected population are key drivers of the level and global variation of the cIFR, and that many countries in Europe, Northern America, and Australia have comparatively high cIFRs as proportionally more older adults are exposed to an infection with COVID-19. The scenarios illustrate that the cIFR would respond differently in each country to changes in age-specific IFRs, with the potential impact being the larger the older the infected individuals are. Age-specific IFRs could be higher due to shortages in critical medical resources or spread of deadlier variants of SARS-CoV-2 (10) and they could be lower as courses of COVID-19 are expected to be less deadly after vaccination (14). However, full vaccination of affected populations and breakthroughs in medical treatment take time (9,13). The sensitivity analysis demonstrates that high-quality input data are essential to accurately estimate the cIFR and that monitoring systems for collecting and preparing COVID-19-related data are worthwhile to be refined worldwide.

## Materials and Methods

Readers find detailed information on the essential materials and extended methods used in this study in the *Supplementary Information*. This GitHub repository: https://github.com/christina-bohk-ewald/assess-total-infection-fatality-burden-of-COVID-19 provides the R-code. Additional information are available in ref. 11.

## Data Availability

Data is available online and can be downloaded using the R-code provided on GitHub: https://github.com/christina-bohk-ewald/assess-total-infection-fatality-burden-of-COVID-19

## Supplementary Information

### Essential datasets

#### Input data from seven different sources

In our study, we estimate the crude infection fatality rate (cIFR; the definition of the cIFR is detailed in a separate section below) based on input data from seven sources. The first data source is the COVerAGE database (1). It provides numbers of reported deaths and confirmed cases attributable to COVID-19 by 5-year age groups (0-4, …, 95+ years old), for both sexes combined and also separately for women and men, for 111 countries on a daily basis since 3 January 2020. To generate the cIFR estimates presented in this study, we have used data from the COVerAGE database (1) as from 14 April 2021.

The second data source is the Short-term Mortality Fluctuations (STMF) database (2). It provides all-cause deaths for 38 countries on a weekly basis. The STMF has data by age groups, standardized and originally reported, for all 38 countries. The length of the time series of weekly deaths differs across the countries covered. To estimate excess deaths attributable to COVID-19 in this study, we have used the STMF input data files as of 15 March 2021 (3).

The third data source are the UN World Population Prospects (UNWPP) (4) that provide demographic data by age and sex for many countries worldwide over time. In this study, we have used UNWPP data on remaining life expectancy by age, population size by age, and population median age for all countries of the COVerAGE database (1) of the calendar year 2019.

The fourth data source are the Spanish population seroprevalence survey data of the Ministerio de Sanidad, Gobierno de España (18) that provides estimates of COVID-19-related infections by sex and 5-year age groups (0-4, …, 90+ years old). These sex-specific COVID-19-related seroprevalence estimates have been collected during four rounds of nationally representative surveys by the Health Ministry of the Spanish government between March and November 2020 and include data of 51 409 participants. The COVID-19 infection prevalence is reported by 5-year age groups and sex, including 95% confidence intervals (5). We have used these data to estimate sex-specific baseline IFRs by age for Spain (6).

The fifth, sixth, and seventh data source are baseline IFRs by 10-year age groups for both sexes combined from (i) China as published in Verity et al. (7), (ii) France as published in Salje et al. (8), and (iii) 28 highly developed countries and regions, summarized in a universal regression function, as published in Levin et al. (9).

#### Code and data availability

The materials and methods used as well as the results generated in this study are open and freely available. Detailed information, including access to and preparation of input data and carrying out the analyses, are given in this Supplementary Information and deposited in this GitHub repository: https://github.com/christina-bohk-ewald/assess-total-infection-fatality-burden-of-COVID-19. This GitHub repository also provides the R-code and regularly updated and extended results. For additional information, see ref. 10.

## Extended Methods

### Overview

We provide full details and descriptions of the estimation procedures used in this study, including the definition and computation of the cIFR, the scaling of baseline IFRs by age, and the estimation of the infected population by age. We also provide technical details and descriptions of empirical results, including the spatio-temporal distribution of the cIFR, the correlation between the cIFR and population median age, the uncertainty estimates of the cIFR, and the scenarios illustrating the impact on the cIFR when changing scaled baseline IFRs by age. We also give detailed information on estimating the cIFR by sex and on conducting sensitivity analyses using cumulative numbers of excess deaths instead of reported deaths and approximating the age structure of infected individuals with the age structure of confirmed cases. We provide links to all the data and R-code that we have used in this study.

### Define the crude infection fatality rate (cIFR)

The cIFR quantifies how fatal an infection with COVID-19 is overall. It is defined as total number of deaths (*D*) over total number of infections (*I*). To estimate the cIFR of a country of interest (COI) at a specific time point *t*, we sum across all ages *x* the product of the scaled infection fatality rate by age 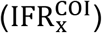 and the age-specific proportion of the infected population 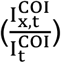 at time *t*:

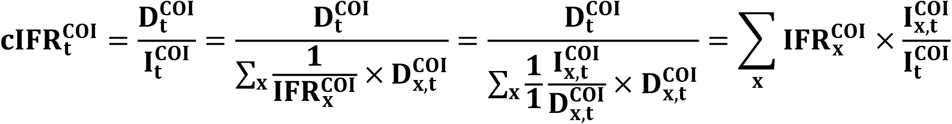

As raw input data we take cumulative numbers of reported deaths by age 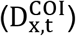 and baseline IFRs by age (IFR_*x*_). We use the demographic scaling model (3) to transform the baseline IFRs by age into country-specific IFRs by age 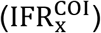 and to estimate the size and age structure of the infected population by age 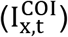 based on these input data. The cumulative deaths are time-variant, the 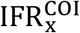 are assumed to be time-invariant. To avoid problems of small numbers, we estimate the cIFR for a country of interest at a specific point in time only when there are at least 200 cumulative deaths.

### Scale baseline IFRs by age for a country of interest

A central input for estimating the cIFR are country-specific IFRs by age (IFR_x^COI). To derive them we scale baseline IFRs by age (5,7,12) from a reference country onto a country of interest via remaining life expectancy (4) using the demographic scaling model (11).

More specifically, we interpolate the broad age schedules of the baseline IFRs (10-year age groups: 0-9, …, 80+) and of remaining life expectancy of abridged life tables of 2019 (5-year age groups: 0-1, 1-4, 5-9,.., 95-99, 100+) onto a fine grid of fractions of single years of age using cubic splines (11). We then assign the same IFR to individuals of the reference country (RC) and the country of interest (COI) that have the same number of life years left 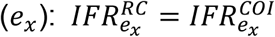. This scaling procedure allows to account for cross-country differences with respect to population age, preconditions, and health care facilities. After this scaling process, we aggregate the scaled baseline IFRs by age for a country of interest from the fine grid into 5-year age groups (0-4, …, 95+) for the subsequent analysis. All data and R-code that we have used to obtain the scaled IFRs by age for both sexes combined for all countries of the COVerAGE database (1) are openly available in the specific version of the GitHub repository as of 16 April 2021: https://github.com/christiandudel/ifr_age/blob/a62a844e0645ecbf4f9c14002648942ca555b1ae/R%20code/IFR_variants.R

We use the same procedure to scale the sex-specific baseline IFRs by age (15) with the demographic scaling model (11). As additional input we take life expectancy of abridged life tables for women and men of 2019 (4). All data and the R-script that we have used to obtain the scaled baseline IFRs for women and men for all countries of the COVerAGE database (1) are openly available in the following version of the GitHub repository as of 16 April 2021: https://github.com/christina-bohk-ewald/assess-total-infection-fatality-burden-of-COVID-19/blob/65d8c99d70cb8b7e0a50f65c7728b66a2b5076f5/step-0-point4-sex_specific_IFRs_selfcontained.R

Additional information (11) shows the level and age profile of the three baseline IFRs for both sexes combined (7-9). All three baseline IFRs predict that infection fatality exponentially increases with age. However, Salje’s baseline IFRs (8) are lowest at medium adult and old ages; Verity’s baseline IFRs (7) are highest at medium adult and old ages, but lowest at oldest old ages; and Levin’s baseline IFRs (9) are lowest at younger ages and highest at oldest old ages. These general differences in the baseline IFRs by age are reflected in the scaled IFRs by age in each country. The additional information (10) also shows scaled IFRs by age for Australia and Japan based on data of Verity et al. (7); more information and empirical examples on scaling baseline IFRs by age are given in (11). The additional information (10) also shows the Spanish baseline IFRs by age and sex; they are consistently lower for females than males at all ages. Since this scaling procedure of baseline IFRs by age requires only little data it is broadly applicable in countries worldwide regardless of whether they have poor or rich data on COVID-19 (11).

### Estimate the size and age structure of the infected population for a country of interest

Another central input for estimating the cIFR is the age structure of the infected population in a country of interest 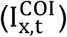. We use the demographic scaling model (10) to estimate it as the sum across all ages *x* of the fraction of cumulative numbers of reported deaths by age at time 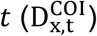 from the COVerAGE database (1) and the scaled IFRs by age 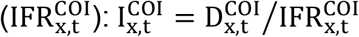. In this study we use the estimated cumulative number of infections by 5-year age groups (0-4, …, 90-95, 95+) and time.

### Spatio-temporal distribution of the cIFR

The spatio-temporal distribution in Fig. 1A of the main text consists of daily cIFR estimates for all countries of the COVerAGE database (1) between 28 March 2020 and 31 March 2021 for which we have at least 200 cumulative deaths. In total, we have 12 086 data points by country and date. We estimate the daily cIFR in Fig. 1A based on scaled baseline IFRs by age of Verity et al. (7) and cumulative numbers of reported deaths by age and time. All data and the R-code that we have used to estimate the cIFRs are openly available in the following version of the GitHub repository as of 16 April 2021: https://github.com/christina-bohk-ewald/assess-total-infection-fatality-burden-of-COVID-19/blob/b2a082b75bf46879472de617d116037cae2f3cd0/step-2-spatio-temporal-dist-total-IFR.R

### Correlation between log of cIFR and population median age

In Fig. 1C of the main text we show the log-linear relationship between the log of all 12 086 cIFR estimates and population median age. Pearson’s r is 0.9. All data and the R-code that we have used to estimate the correlation between the log of cIFR and population median age are openly available in the following version of the GitHub repository as of 16 April 2021: https://github.com/christina-bohk-ewald/assess-total-infection-fatality-burden-of-COVID-19/blob/b2a082b75bf46879472de617d116037cae2f3cd0/step-3-corr-btwn-IFR-age-P-I.R The additional information (10) shows the correlation between log of all 12 086 cIFR estimates and median age of infected population. Pearson’s r is 0.9. However, possible circular effects and both quantities having been computed using COVID-19-related deaths may make these results statistically less valid. The additional information (10) also shows the correlation between log of all 7 909 cIFR estimates (based on the age structure of confirmed cases rather than of estimated infections) and median age of confirmed cases from the COVerAGE database (1). Pearson’s r is 0.9. However, the number of confirmed cases underestimates the number of infections due to a lack of reliable COVID-19 data (11,12). Despite these limitations (partly statistical and data-related), these correlation analyses suggest that the age structure of the general and infected population are key drivers of the level and global variation of the cIFR.

### Uncertainty estimates of the cIFR

In Fig. 1, panels D through G, we show the 95% prediction intervals for the latest cIFR estimate as of 31 March 2021. Following the analysis of the demographic scaling model (11), we arrive at these uncertainty estimates by using the lower and upper boundary of the 95% credible interval of the corresponding baseline IFRs by age (7-9), which have been derived through Bayesian analysis. We insert these lower and upper boundaries of the baseline IFRs by age in the formulas for estimating the size and age structure of the infected population and finally the cIFR in order to get their 95% prediction intervals. All data and the R-code that we have used to estimate the 95% prediction intervals of the cIFR are openly available in the following version of the GitHub repository as of 16 April 2021: https://github.com/christina-bohk-ewald/assess-total-infection-fatality-burden-of-COVID-19/blob/b2a082b75bf46879472de617d116037cae2f3cd0/step-2-spatio-temporal-dist-total-IFR.R

### Scenarios of the latest cIFR

Figure 1, panels E through G, show how the original cIFR estimates as of 31 March 2021 would change if the scaled IFRs by age would be different as a result of, e.g., improved medical treatment (13,14), broad access to vaccination (15), or increased spread of deadlier SARS-CoV-2 variants (16), or potential misestimation. To simulate potential changes in the scaled IFRs by age we multiply them with a proportional factor: 0.4, 0.6, 0.8, 1, 1.2, 1.4, and 1.6. We then use these modified IFRs by age to re-estimate the cIFR and analyze their difference to the original cIFR estimates. We do this analysis for three baseline IFRs by age (7-9) and for all countries of the COVerAGE database (1) for which we have at least 200 cumulative deaths as of 31 March 2021. All data and the R-code used to generate and analyze these cIFR scenarios are openly available in the following version of the GitHub repository as of 16 April 2021: https://github.com/christina-bohk-ewald/assess-total-infection-fatality-burden-of-COVID-19/blob/b2a082b75bf46879472de617d116037cae2f3cd0/step-5-scenarios-IFR.R

### Estimate the cIFR by sex

We estimate the cIFR separately for women and men with sex-specific input data. That is, we use sex-specific baseline IFRs by age of Spain provided by Acosta (6) and cumulative numbers of sex-specific deaths by age reported to the COVerAGE database (1). We then analyze the difference between female and male cIFRs in Fig. 2A of the main text.

We first estimate the Spanish baseline IFRs for women and men by 5-year age groups based on sex-specific numbers of deaths and infections in four steps. In a first step we collect COVID-19-related deaths by age. For ages 0 through 64 years old, we take sex-specific deaths that have been reported to the COVerAGE database (1). For ages 65 through 90+ years old, we use excess deaths by sex that we have estimated based on weekly deaths of the STMF input database for Spain (2). (We provide more details on estimating excess deaths using STMF data in a separate section below.) In a second step, we obtain estimates of the number of infection by sex and 5-year age groups, 0 through 90+ years old, from the four rounds of the Spanish nationally representative seroprevalence study (ENE-COVID) (5). In a third step, we then estimate the baseline IFRs by age as the fraction of sex-specific deaths by age over sex-specific infections by age. In a fourth step, we also estimate 95% confidence intervals for these Spanish baseline IFRs by age for women and men using the uncertainty estimates reported in the prevalence survey (5). All data and the R-code that we have used to obtain these Spanish baseline IFRs by age for women and men (6) are openly available in the following version of the GitHub repository as of 15 March 2021: https://github.com/kikeacosta/ifr_age_spain/tree/e32df2f7fc64939ab32e97d75ff6 25bda8af24f8

We then continue with scaling these Spanish baseline IFRs by age and sex (6) for a country of interest via remaining life expectancy of abridged life tables by sex of 2019 (4) using the demographic scaling model (11). And we take cumulative numbers of sex-specific deaths by age for a country of interest reported to the COVerAGE database (1). Based on these sex-specific input data, cumulative deaths by age and sex and scaled IFRs by age and sex, we estimate the infected population by age and sex and finally the cIFR by sex in a country of interest at a specific point in time. (We provide details on scaling baseline IFRs by age and estimating the infected population by age in separate sections above.)

In total, we have 7 370 data points for each sex to calculate the difference between female and male cIFRs for the same country and date. All data and the R-code that we have used to estimate cIFR by sex are openly available in the following version of the GitHub repository as of 16 April 2021: https://github.com/christina-bohk-ewald/assess-total-infection-fatality-burden-of-COVID-19/blob/b2a082b75bf46879472de617d116037cae2f3cd0/step-6-total-IFR-by-sex.R

### Estimate the cIFR based on excess deaths

We analyze the sensitivity of the cIFR with respect to estimating the age structure of the infected population based on reported deaths or excess deaths.

We first estimate excess deaths by 5-year age groups for both sexes combined based on weekly all-cause mortality data of the STMF database (2) in four steps. We define excess mortality as the difference between observed mortality and baseline mortality that is equal to the mortality expected in the absence of the coronavirus pandemic. In a first step, we collect weekly all-cause mortality data for 21 countries of the STMF database (2) as well as for Mexico and Peru from their national vital statistics systems (18-20). In a second step, we collect and interpolate annual population counts for the same countries of the Human Mortality Database (HMD; 21) and the UNWPP (4) in order to get weekly exposures. In a third step, we estimate the mortality baseline by fitting a GLM model with a Poisson distribution, using weekly deaths as response variable and weekly exposures as offset, between 2010 and 2019. We exclude data of winter weeks in order to avoid mortality disturbances that have been caused by seasonal influenza epidemics. We also estimate 95% prediction intervals for the mortality baseline based on 2 000 bootstrapping iterations. In a fourth step, we finally compute excess mortality as the difference between observed mortality and expected or baseline mortality. We do this only for those weeks for which we find that observed mortality exceeds the upper border of the 95% prediction interval of the baseline mortality. All data and the R-code used to estimate excess mortality are openly available in the following version of the GitHub repository as of 15 March 2021: https://github.com/kikeacosta/excess_mortality/tree/bad697002f759a642efcfdebd dd0498c4e9bdf2a

We then continue with scaling the baseline IFRs by age of Verity et al. (7) for a country of interest via remaining life expectancy of abridged life tables of 2019 using the demographic scaling model (3). And we then estimate the age structure of the infected population using either cumulative numbers of deaths by age as reported to the COVerAGE database (1) or, alternatively, the excess deaths by age that we have estimated in the previous step. (We provide details on scaling baseline IFRs by age and estimating the infected population by age in separate sections above.) We then estimate the daily cIFR using the scaled IFRs by age and the age structure of the infected population based on either reported deaths or excess deaths.

The additional information (10) shows the difference between these two cIFR estimates in order to analyze the sensitivity of the cIFR with respect to estimating the age structure of the infected population based on reported deaths or excess deaths. In total, we have 772 data points for the same country and date. We find the cIFR to be sensitive towards the source of deaths by age. The absence of a systematic pattern in these cIFR differences in addition to uncertainty in estimating excess deaths point to quality differences not only in the source of input data (please see the separate section on quality of input data from different sources below), but also in the resulting cIFR estimates. In consequence, we recommend to use reported deaths instead of excess deaths when estimating the cIFR. All data and the R-script used to estimate these two variants of the cIFR based on reported deaths or excess deaths are openly available in the following version of the GitHub repository as of 16 April 2021: https://github.com/christina-bohk-ewald/assess-total-infection-fatality-burden-of-COVID-19/blob/b2a082b75bf46879472de617d116037cae2f3cd0/step-7-sensitivity-IFR.R

### Estimate the cIFR based on the age structure of confirmed cases

We analyze the sensitivity of the cIFR with respect to approximating the age structure of infected individuals with the age structure of confirmed cases.

In a first step, we scale the reference IFRs by 5-year age groups of Verity et al. (7) for a country of interest via remaining life expectancy of abridged life tables of 2019 (4) using the demographic scaling model (11). (We provide details on scaling baseline IFRs by age in a separate section above.)

In a second step, we estimate the age structure of the infected population based on cumulative numbers of deaths by 5-year age groups that have been reported to the COVerAGE database (1) and scaled reference IFRs by age. (We provide details on estimating the infected population by age in a separate section above.) As an alternative, we approximate this estimated age structure of the infected population with the observed age structure of confirmed cases from the COVerAGE database (1).

In a third step, we estimate the daily cIFR using as input the scaled reference IFRs by age and the age structure of infected individuals or the age structure of confirmed cases.

The additional information (10) also shows the difference between these two cIFR estimates in order to analyze the sensitivity of the cIFR with respect to approximating the age structure of infected individuals with the age structure of confirmed cases. In total, we have 8 429 data points for the same country and date. We find the cIFR to be sensitive towards the source of the age structure of infections. More specifically, the absence of a systematic pattern in these cIFR differences in addition to cIFR misspecification when using confirmed cases point to quality differences not only in the source of input data (please see the separate section on quality of input data from different sources below), but also in the resulting cIFR estimates. In consequence, we recommend to use the age structure of the infected population instead of the age structure of confirmed cases. All data and the R-code used to estimate cIFR based on the age structure of infected individuals and of confirmed cases are openly available in the following version of the GitHub repository as of 16 April 2021: https://github.com/christina-bohk-ewald/assess-total-infection-fatality-burden-of-COVID-19/blob/b2a082b75bf46879472de617d116037cae2f3cd0/step-7-sensitivity-IFR.R

### Quality of input data from different sources

A closer look at the results of the sensitivity analysis indicates that the differences in cIFR based on different input data vary considerably across countries—a pattern that can perhaps be explained with small numbers, but also with potential inconsistencies in the practices adopted for monitoring COVID-19-related data in each country. That is why we provide some more information on the quality of the input data from different sources that we have used in this study to estimate the cIFR.

The COVerAGE database (1) contains high-quality data on confirmed cases and deaths by age and sex attributable to COVID-19 that are extracted from reports published by official governmental institutions worldwide. Besides data in the originally reported age bands, the COVerAGE database (1) offers two additional files with data harmonized in 5- and 10-year age groups. The complete details on all steps of collecting and preparing the data (e.g., age harmonization) are consigned in the COVerAGE database Method Protocol (21). The quality of the underlying data on confirmed COVID-19-related cases and deaths included in the COVerAGE database (1) is heterogeneous and difficult to evaluate. This stems primarily from inconsistent definitions and testing strategies within and across populations (1,11,12). To facilitate a cautious approach regarding data interpretation and data limitations, the COVerAGE database (1) offers additional information on sources, metadata, and quality for each country. An up-to-date list of original data sources can be found in the project repository at: https://timriffe.github.io/covid_age/DataSources.html. The metadata of the database also include definitions of confirmed COVID-19-related cases and deaths. Finally, the COVerAGE database (1) also provides a quality metrics object (1,22; https://osf.io/qpfw5/) that includes country-specific indicators on age reporting completeness, degree of age harmonization, complementary measures on testing, and comparisons of total numbers of confirmed cases and deaths with data from the Our World in Data database (23).

In general, we expect the STMF data series (17) to provide high-quality data on weekly all-cause mortality, because it includes data only for countries where the census and vital registration system cover close to 100 percent of the general population. Nevertheless, the quality of this STMF data can be compromised for two main reasons. First, there are differences across countries with respect to the date of death definition. In some countries the date of death corresponds to the occurrence of the death and in other countries to the registration of the death. Second, due to delayed registration of deaths, the data corresponding to the last weeks of a calendar year as well as data for all weeks of the most recent calendar year might be incomplete. The STMF does not perform any adjustments for the above mentioned potential data quality problems. More country-specific information regarding data formats and adopted methods are provided in a preliminary note on the STMF (https://www.mortality.org/Public/STMF_DOC/STMFNote.pdf) as well as in corresponding metadata (https://www.mortality.org/Public/STMF_DOC/STMFmetadata.pdf).

We expect the population level survey data on the prevalence of antibodies against SARS-CoV-2 among Spanish women and men to be of high quality. The four rounds of these nationally representative seroprevalence surveys were conducted between April and November 2020 (5). As the initial survey was performed one month after the peak incidence of COVID-19 on 20 March 2020, decreases of antibodies after an infection are not expected to affect the results of these prevalence estimates (24).

